# Data rectification to account for delays in reporting disease incidence with an application to forecasting COVID-19 cases

**DOI:** 10.1101/2024.04.08.24305398

**Authors:** Yunus A. Abdulhameed, Samuel Roberts, Jacob B. Aguilar, James Kercheville, Juan B. Gutierrez

## Abstract

Effective monitoring of infectious disease incidence remains a major challenge to public health. Difficulties in estimating the trends in disease incidence arise mainly from the time delay between case diagnosis and the reporting of cases to public health databases. However, predictive models usually assume that public data sets faithfully reflect the state of disease transmission. In this paper, we study the effect of delayed case reporting by comparing data reported by the Johns Hopkins Coronavirus Resource Center (CRC) with that of the raw clinical data collected from the San Antonio Metro Health District (SAMHD), San Antonio, Texas. An insight on the subtle effect that such reporting errors potentially have on predictive modeling is presented. We use an exponential distribution model for the regression analysis of the reporting delay. The proposed model for correcting reporting delays was applied to our recently developed SEYAR (Susceptible, Exposed, Symptomatic, Asymptomatic, Recovered) dynamical model for COVID-19 transmission dynamics. Employing data from SAMHD, we demonstrate that the forecasting ability of the SEYAR model is substantially improved when the rectified reporting obtained from our proposed model is utilized. The methods and findings demonstrated in this work have ample applicability in the forecasting of infectious disease outbreaks. Our findings suggest that failure to consider reporting delays in surveillance data can significantly alter forecasts.

## 1 Background

In December of 2019, a novel coronavirus (SARS-CoV-2) was first reported in the City of Wuhan, Hubei Province, China. On January 30^th^, 2020, the SARS-CoV-2 outbreak was declared a public health emergency of international concern. On March 11^th^, 2020 the World Health Organization declared a global pandemic (*43*).

Early in the pandemic, two resources rose to prominence as sources of data. The Coronavirus Resource Center (CRC) at Johns Hopkins University (*11*), and The New York Times (NYT) coronavirus database (*50*). An important characteristic of these privately-maintained public databases is that after data is initially entered, it is usually not updated.

Various models have been used during the COVID-19 outbreak. Applications of these models include to inform public health policies (*13,36*), to assess the impact of government interventions (*5, 31, 45*), to project hospital utilization (*38*), and to assess disease transmission dynamics (*33*). The outcomes of these models rely on three important factors: (i) the quality of the case data used to calibrate the model, (ii) the validity of parametric assumptions, and (iii) the number of parameters. The current focus on improving predictive case modeling has been centered around the limitations that arise from poor parametric assumptions, and the large number of parameters used in some models. As a result, the problem of poor data quality has received comparatively little attention.

Quality epidemiological data is central to infectious disease surveillance and modeling. From a public health viewpoint, accurately monitored data is a crucial resource for understanding the true extent of population-level disease progression during an epidemic event (*17*). This knowledge can then be used to support critical decision making by public health authorities. Previous epidemics such as Zika, Ebola and Swine flu have revealed the usefulness of accurate epidemiological data for emergency preparedness (*9*), vaccine distribution (*22*), and planning for the future demand of critical infrastructure (*30, 41*). From an infectious disease modelling viewpoint, quality data is critical to ensuring accurate epidemic forecasting (*39*). Furthermore, the accuracy of epidemiological parameters required by compartmentalized models (*28*), agent based models (*8, 15, 24*), and/or training and validation of machine learning and statistical models (*21,48,52*) is directly related to the quality of input data used to estimate these parameters.

Data quality is affected by case tracing and delays in reporting. For case counts, the *epidemiological event date*, *E*, is often defined in practice as: (i) *E* = Date of onset of symptoms, (ii) if date of onset is not available, then *E* = date of sample collection, (iii) if date of sample collection is not available, then *E* = date of lab report, and (iv) if date of lab report is not available, then *E* = date entered into database.

The epidemiological event date changes continuously as case trace investigations take place, which can last a variable number of days depending on the operations of the municipality collecting the data. As a result, the case counts for past dates become updated with the results obtained from the case trace investigations, with older unresolved cases being dropped from the tracing process. Another source of variation in case reporting is the generalized inherent delay due to the time it takes for samples to be analyzed, the time to register an identified case into a database, and overburdened surveillance systems sometimes operating with a throughput lower than the influx of new cases. It is important to note that the CRC and NYT databases do not change the data once it is reported. This raises concerns in relation to the accuracy of predictive models since many modelers do not have access to raw case tracing data.

The challenge of reporting delays was first studied by Harris (*26*) who contrived the problem as a partial multinomial distribution. Thereafter, various statistical methods (*6,49,55*) have been employed to address the problem of delays from infection to symptom onset, as well as the subsequent delays that arise before the data is entered into surveillance databases. These techniques broadly fall into parametric and non-parametric methods. The parametric approach accounts for reporting delay based on the assumption that delayed case data belongs to a parametric family of probability distributions. A number of parametric-based methods that model the reporting delay in previous disease outbreaks (*6, 10, 19, 27*), and in the COVID-19 pandemic (*1, 37*) have been reported.

The non-parametric approach provides an estimate of the reporting delay distribution without making any parametric assumption about the form of the underlying delay distribution. There are two common computational methods employed for finding non-parametric estimates of the reporting delay distribution. The first is based on the generalized linear model (e.g. Poisson regression or the non-parametric back-propagation of delayed cases) which requires cross-classification of reported cases by calendar time of diagnosis and reporting delays (*6, 55*). The back-propagation process uses a diagnosis distribution to estimate the number of case counts for previous time periods (*4, 32, 34*), and has recently been used to assess infection incidence of COVID-19 (*37*). The second method employs a survival analysis approach which involves expressing the delay distribution as a product of conditional probabilities, from which an estimate for the reporting delays is obtained.

Another approach used to account for delays in case data produces real-time estimates for the current number of confirmed infections while correcting for underreporting. This new technique is referred to as ‘nowcasting’ (*18, 20, 42,51, 54*). Depending on the assumptions it is premised on, this method could be considered either a parametric or a nonparametric approach. For example, nowcasting procedures based on the non-parametric approach have been used to assess the Shiga toxin–producing E. coli (STEC) O104:H4 outbreak in Germany (*29*). On the other hand, to study influenza A/H1N1 during the 2009 pandemic, a nowcasting procedure that involved the parametric method was utilized (*12*). In addition, nowcasting has been used for real-time COVID-19 tracking (*23, 25, 47*).

Data assimilation techniques have also been used (*14,16,40*) to study reporting delays. These methods begin with a wide prior distribution for the model parameters from which a posterior estimation of parameters leads to model predictions that closely agree with the observations. An interesting example of data assimilation is Abott et al. (*1*), since there is explicit consideration of reporting delay. In all these methods, model updates occur when new observations become available.

In this present study, we consider an approach that is notably different from nowcasting and data assimilation. It is informed by the comparison of the totality of records of daily cases from public databases against the real-time number of hospitalizations and cases. The COVID-19 data used here is for the City of San Antonio, Texas, the seventh largest city in the US, with a population of 1.5 million people. It is the most visited city in Texas, and the 17*^th^* most visited city in the US, attracting 37 million visitors in 2019 (*3*). Given the high density of visitors year-round, the city has the potential to become an epicenter of transmission during a pandemic. The City of San Antonio entered a partnership with Bexar County to create the San Antonio Metro Health District (SAMHD); this entity is in charge of collecting case data for infectious diseases.

To better account for delays in reporting COVID-19 case-data, and to accurately forecast the number of confirmed cases, we adopted the following approach. First, we analysed the complete set of epidemiological event dates for confirmed COVID-19 cases and developed an algorithm that rectifies delays in public epidemiological data. Then, we incorporated the proposed rectification algorithm into our compartmentalized SEYAR model to project the number of cases in Bexar County, Texas, whilst taking into account reporting delays.

## 2 Methods

### 2.1 Epidemiological data extraction

We employed two different data sets which were obtained from two different sources. The first being the COVID-19 case-data retrieved from the CRC. The second was the data regarding the epidemiological date for COVID-19 cases acquired from the SAMHD between April 4, 2020 through June 28, 2020. The epidemiological event dates received daily from the SAMHD include; (i) illness onset, (ii) sample collection, (iii) test result, and (iv) case entered into the database. It is usually not the case that information is available for every epidemiological date mentioned ((i)-(iv)); but at least one of these dates is available for each confirmed case.

### 2.2 Extracting the delay distribution from surveillance data

Let *a_jk_* be the data reported on day *k* into the SAMHD registry, for each day *j* subject to 1 *≤ j ≤ k*. Let *b_jk_* be the ‘stable’ data where ‘stable’ means that as time progresses, the number of cases for day *b_jk_* does not change. The residue *r_jk_* = *a_jk_ − b_jk_* is modeled as an exponential distribution characterized by *y_j_* = *pe^qj^* where the parameters *p* and *q* denote the correction rate.

The least-squares objective function is expressed as

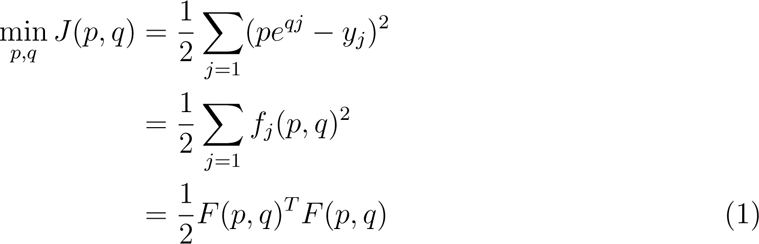

where *F* is the vector-valued function *F* (*p, q*) = (*f*_1_(*p, q*) *f*_2_(*p, q*) *… f_i_*(*p, q*))*^T^*. The derivatives are made less cluttered by scaling the problem by 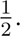 The gradient of *f* is

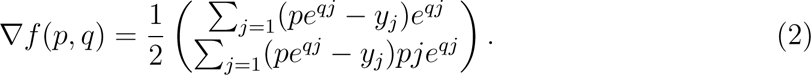

Suppose the solution of the least-squares problem is given by *p_∗_*, *q_∗_*, then *f* (*p_∗_,*q*_∗_*) = 0. This implies that for all *j*, *f_j_*(*p_∗_, q_∗_*) = 0, suggesting that the model is in agreement with the data with minimal error. Consequently, *F* (*p_∗_, q_∗_*) = 0 for *p ≈ p_∗_*, *q ≈ q_∗_* which justifies that the required first-order condition is met.

After training, the obtained parameters were averaged and used to test the model. Finally, the averaged estimated parameters obtained from the model validation process were employed to estimate the delayed daily case counts for each data set reported by the CRC.

### 2.3 Minimizing the time lag between CRC and SAMHD data

The number of confirmed cases for Bexar County entered into the SAMHD registry was updated each day to reflect the number of delayed cases, hence providing an accurate baseline. Our objective is to minimize the error between the data entered into the SAMHD and CRC registries.

Let *a_mk_*and *c_nk_*be the case data reported into the SAMHD and CRC registries respectively on day *k*, for each day *m* (for SAMHD data) and *n* (for CRC data) subject to 1 *≤ m ≤ k* and 1 *≤ n ≤ k* respectively. The optimal data time Δ*t* that minimizes the time lag between these two case data is estimated by taking the difference between the dates at which the number of cases in *a_mk_* and *c_nk_* are equal. The optimal data time objective function is an average expressed as

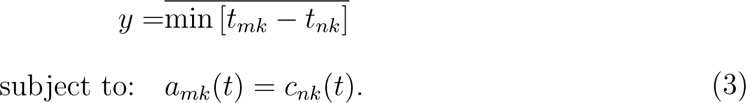

Finally, 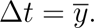

### 2.4 SEYAR Model Modification

Consider the *SEYAR* dynamical system (4) introduced by Aguilar et. al (*2*), which describes the dynamics of COVID-19 transmission in a human population by decomposing the total host population (*N*) into the following five epidemiological classes: susceptible human (*S*), exposed (*E*), symptomatic (*Y*), asymptomatic (*A*), and recovered (*R*).

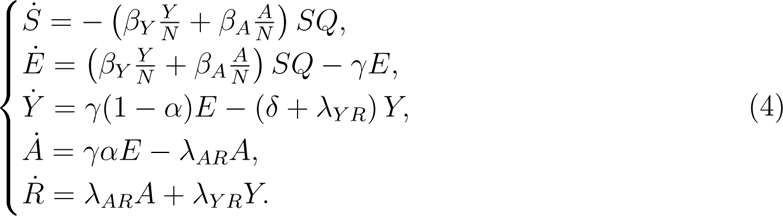

Here *β_Y_*, *β_A_* denote the effective contact rates for symptomatic and asymptomatic carriers respectively, *γ* represents the latent period, 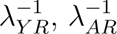 represent the infectious periods for the symptomatic and asymptomatic sub-populations, and *α*, (1 *− α*) represent the probability of becoming asymptomatic and symptomatic upon infection respectively. Moreover, the risk presented at time *t* is represented by *Q*(*t*) = *e^−kt^* and is formulated under the assumption that the rate of change of risk decreases proportionally to the amount of risk present. In the presence of reporting delay, the amount of risk at any time *t* is proportional to the corrected number of reported COVID-19 cases, i.e. the number of corrected cases increases with an increase in the risk of infection. Thus, we arrive at the following modification of (4):

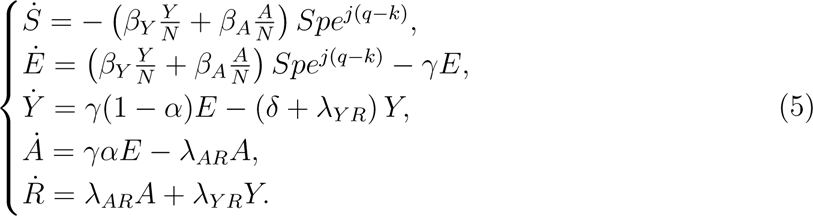

where the parameters *p* and *q* were fitted for a given value of the coefficient of risk mitigation *k*.

To compute the confidence intervals, a sum of 100 bootstrap replications of the case time series were used to find the possible value of the parameter that is close to a global minimum. With the parameters obtained from bootstrapping, the SEYAR model was computed using both datasets. When computing, the model considered the past two weeks of data from the most recent date of available case data.

## 3 Results

Figure 1 summarizes the daily number of confirmed cases in Bexar County, Texas due to the COVID-19 pandemic, as reported by both the SAMHD and CRC. The various solid curves illustrate the daily numbers of cases reported by SAMHD between May 4^th^ 2020 through July 4^th^ 2020. In particular, each solid curve represents a corrected version of the number of cases reported in the past. The dashed curve represents the data reported by the CRC between May 4^th^ 2020 through July 4^th^ 2020. In contrast to the SAMHD data sets, the CRC data for the number of cases reported in the past are not corrected. Moreover, the time lag between the solid and dashed curves was computationally estimated at 8 days.

**Figure 1:**
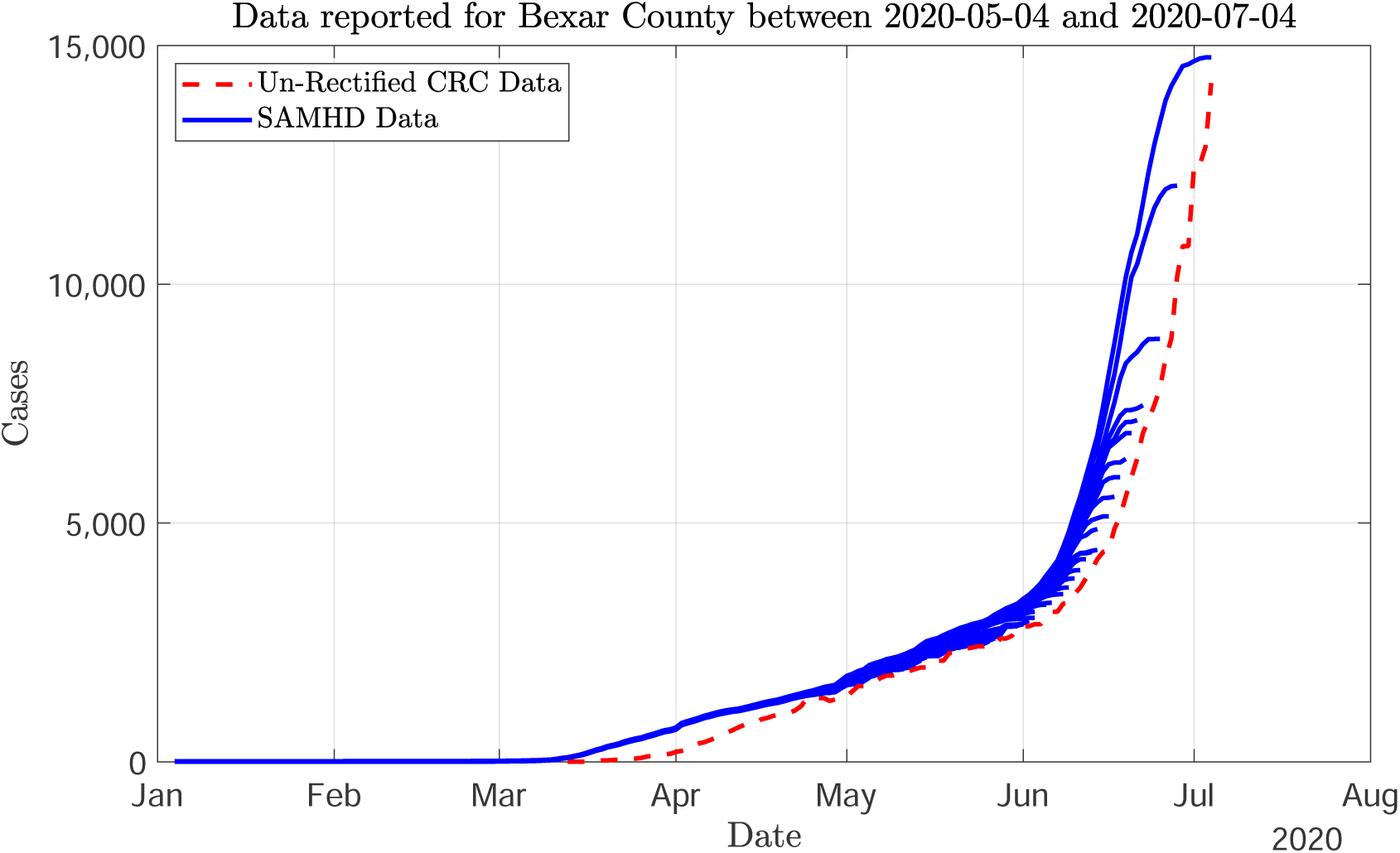
Daily number of confirmed COVID-19 cases in Bexar County, Texas reported for the period between May 4^th^ 2020 through July 4th 2020. Comparison of daily case counts reported by the San Antonio Metro Health District (SAMHD) and the Johns Hopkins Coronavirus Resource Center (CRC). The solid curves represent individual SAMHD data sets across this time period for which previously reported data sets are corrected. The dashed curve refers to the CRC data set for which no corrections are made in the data sets reported in the past.

In Figure 2, the CRC data sets reported between May 4^th^ 2020 and July 4^th^ 2020 were rectified using our proposed data rectification algorithm which was subsequently compared with the baseline SAMHD data set reported on July 4^th^ and the individual unrectified CRC data sets. The Figure indicates congruence between the data transmitted on the same date by the SAMHD, and the rectified CRC data. Additionally, it can be observed that the SAMHD and the CRC reported different numbers of cases for the majority of dates. A significant increase in reporting delay of daily case counts is noticed from approximately the beginning of June and through the conclusion of this study.

**Figure 2:**
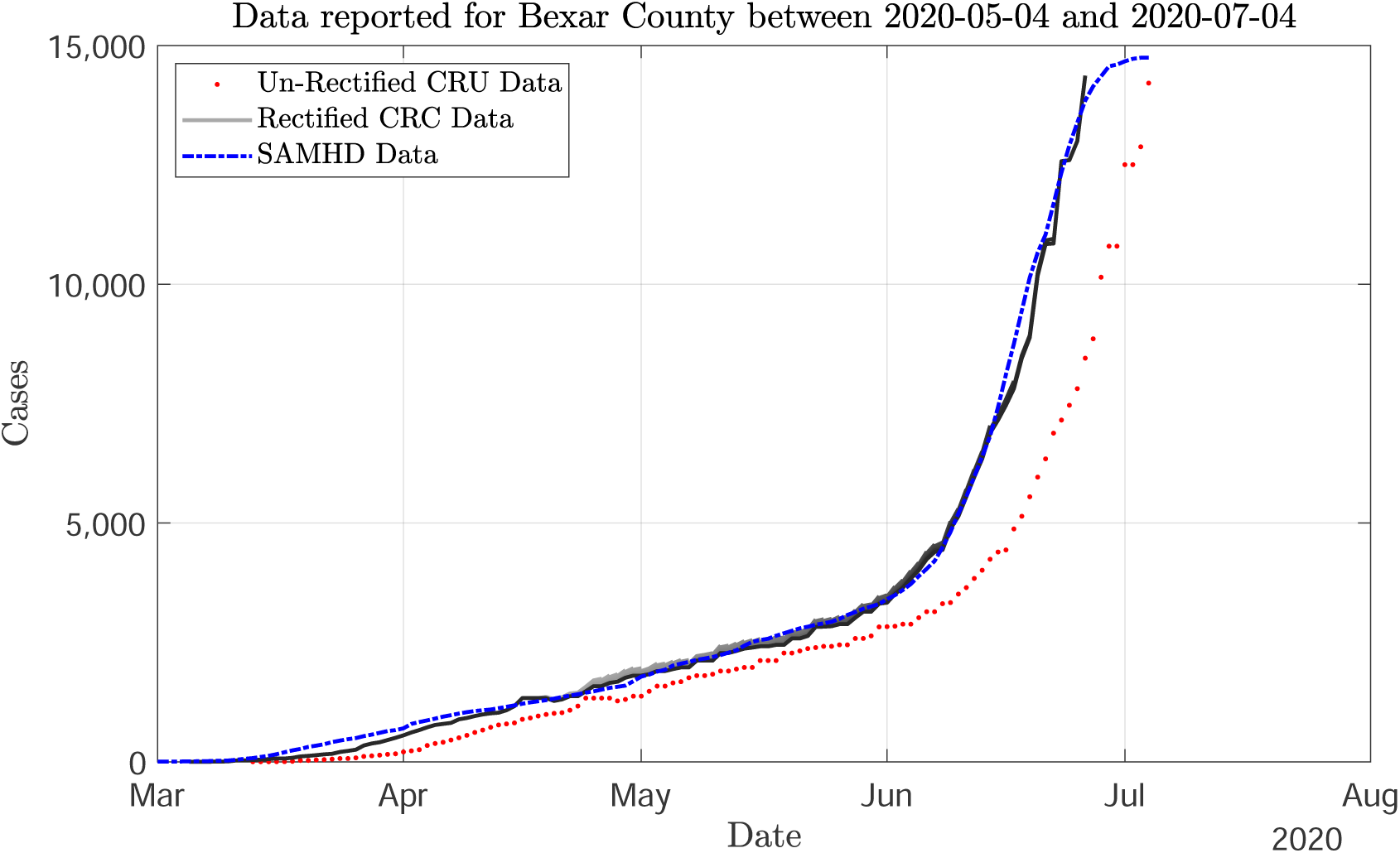
Rectification for individual CRC data sets reported between May 4^th^ 2020 through July 4^th^, 2020. The solid curve indicate the rectified daily confirmed cases that were reported during the pandemic. The dash-dotted curve represents our baseline data, i.e the data set reported by the SAMHD on July 4^th^ 2020. The dotted curve represents the un-rectified CRC data between May 4^th^ 2020 through July 4^th^ 2020.

Taking into account the parameters reported in Table 1, and the five major government mitigation policies that were enacted in Bexar county between March 2020 and July 2020, Figure 3 summarizes an implementation of a SEYAR model calibrated with three sets of data: the time series obtained from CRC, the rectified CRC data, and the data collected from SAMHD. Figure 3 (a) shows a comparison between the average SEYAR model output calibrated using the un-rectified CRC data set reported on June 30^th^ 2020 and the data retrospectively obtained for the month of October 2020. Figure 3 (b) shows a comparison between the average SEYAR model output calibrated using the rectified CRC data set reported on June 30^th^ 2020 and the data retrospectively obtained for the month of October 2020. Figure 3 (c) shows a comparison between the average SEYAR model output calibrated using the SAMHD data set reported on June 30^th^ 2020 and the data retrospectively obtained for the month of October 2020.

**Figure 3:**
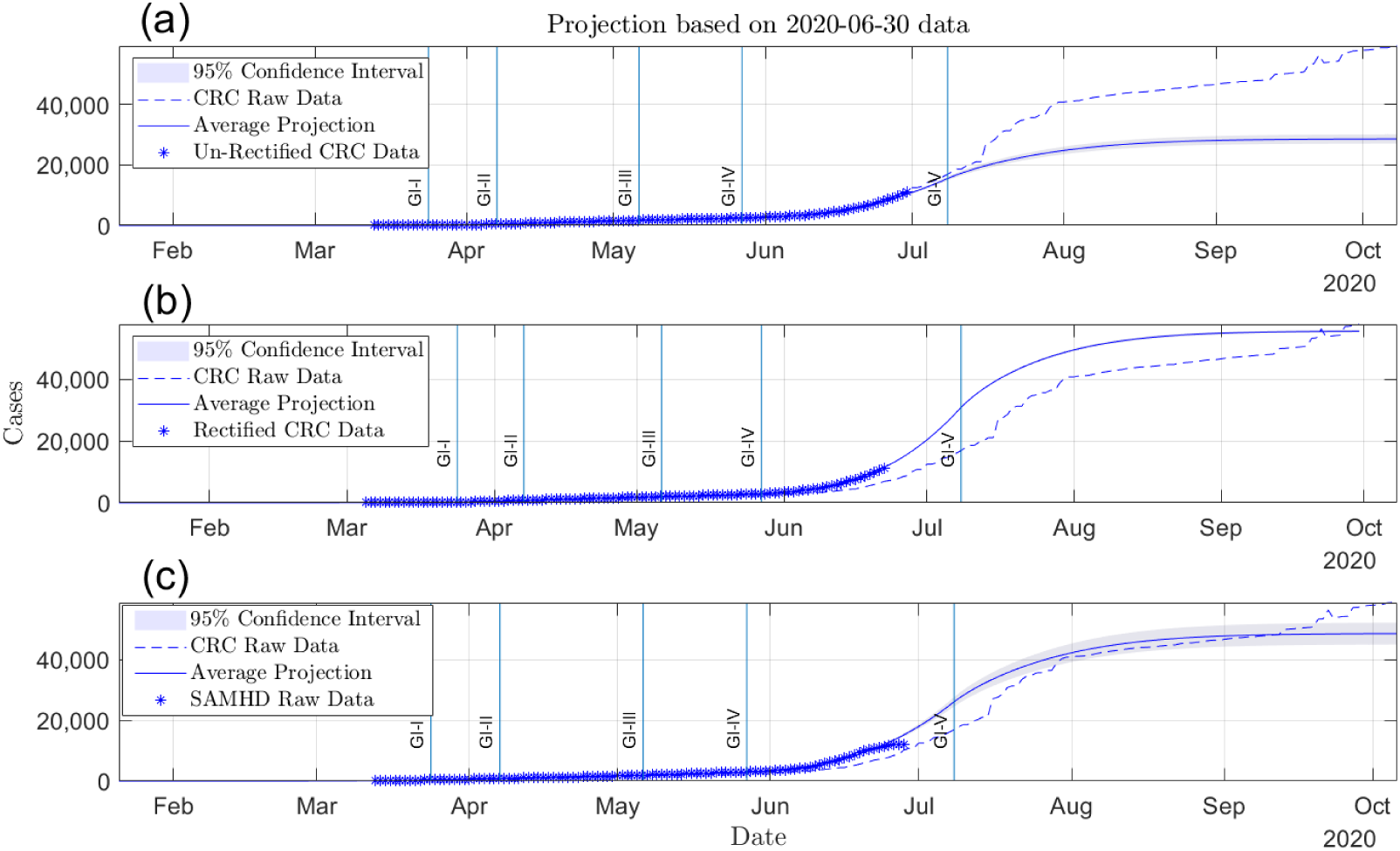
Projections of COVID-19 cases in Bexar County from July 2020 to October 2020, using a SEYAR model calibrated with three data sets: (a) the time series obtained from CRC, (b) the rectified CRC data, and (c) the data collected from SAMHD. The projected number of cases with 95% confidence interval for each of the data sets were obtained under the conditions that applied on July 4^th^ 2020. The blue-dashed line (CRC raw data) denotes the un-rectified daily number of cases reported by the Johns Hopkins Coronavirus Resource Center. The solid vertical lines indicate the timings of various government intervention (GI) strategies.

**Table 1:** Average values of parameters used to compute Figure 3. The fitted parameters were selected from the optimal fit of the model calibrated for Bexar County.

| Parameter | Description | Dimension | Value |  |
| --- | --- | --- | --- | --- |
| *FITTED* |  |  |  |  |
| $\beta_Y$ | Effective contact rate from symptomatic to susceptible. | days<sup>-1</sup> | 1.17 | |
| $\beta_A$ | Effective contact rate from asymptomatic to susceptible. | days<sup>-1</sup> | 1.16 | |
| $k$ | Risk mitigation coefficient | n/a | 0.03 | |
| $n$ | Days prior, patient zero | n/a | 25 | |
| Parameter | Description | Dimension | Mean | Variance |
| *BIOLOGICAL* |  |  |  |  |
| $\gamma^{-1}$ | Mean latent period. | days | 2.99 | 1.36 |
| $\alpha$ | Probability of becoming asymptomatic upon infection. | n/a | 0.21 | 1.48 |
| $\lambda_{YR}^{-1}$ | Mean symptomatic infectious period. | days | 13.5 | 31.8 |
| $\lambda_{AR}^{-1}$ | Mean asymptomatic infectious period. | days | 11.2 | 16 |
| $\delta$ | Disease-induced death rate. | days<sup>-1</sup> | 0.026 | 0.094 |

## 4 Discussion

By analyzing the COVID-19 case count data for Bexar County, Texas, obtained from two different sources –the SAMHD and CRC, we have been able to study how reporting delays in CRC case-data could affect the predictability of trends in the number of daily confirmed cases, as well as the quality of data on which predictive models are calibrated. Accounting for reporting delays in epidemic data enhances the ability to gauge the actual daily case counts for forecasting reliable trends in the number of confirmed cases. This is supported by the findings from the works of Brookmeyer and Liao (*7*), White and Pagano (*53*).

Many of the aforementioned studies on reporting delay rely on an underlying assumed probability density function such as Poisson, Binomial etc. Whilst there are theoretical basis for making such assumptions, unless the chosen distribution is measured from sample data, it remains an assumption which has the potential for introducing a bias. Instead of trying to fit parameters for assumed distributions that make a model fit the data, this present study fits a regression from the complete analysed case tracing data, making the minimum number of assumptions possible. By considering past dates of the pandemic, we can retrospectively evaluate the accuracy of our rectification and forecasting estimates for the number of confirmed cases in Bexar County.

The data pattern that emerged in San Antonio is captured in Figure 1. The solid curves represent cases reported by SAMHD in multiple days. The dashed curve represents data reported by CRC, a source that never updates its initial data entries. It is evident from Figure 1 that case records are constantly corrected at SAMHD by case tracing, and they tend to mimic daily CRC counts with a delay. If data obtained from public databases is substantially different from the true count of cases, then this begs the question of how to create accurate predictive models when most modelers have access only to public databases.

Under-reporting of confirmed cases poses a potentially major challenge in COVID-19 surveillance. Early investigations suggested that most cases are not reported to the Centers for Disease Control (*44*). This justifies our analysis of the cases reported by the CRC for Bexar County (Figure 2), which indicates an incomplete number of daily cases reported. More recent findings indicated a detection rate of only 1–2 % of total actual COVID-19 cases (*35*). As revealed by our analysis of reporting delays, the rate of unreported cases may vary depending on the time lapse between identification of a case and reporting to public registries.

The significant differences observed in the daily number of confirmed cases reported to the CRC is related to the effect of variation in the chosen epidemiological event date caused by case tracing. As a consequence of overwhelmed surveillance systems during the pandemic, there was an usually long turnaround time between the date of onset of illness, date of diagnosis, date of laboratory sample collection, laboratory test result date, and the date that a confirmed case is entered into an official database. These delays in processing lead to imprecise day-to-day case reporting. These challenges have also been shown to introduce a bias in the estimate of case fatality ratio (*46*).

The minimization of the time lag from the reporting of cases and the subsequent adjustment of daily case counts using our proposed method produced a rectification of the public CRC data that approximated data in the official database after case tracing corrections. In terms of rectification of delayed cases, the exponential distribution has provided an accurate model for adjusting delay in case reporting. The agreement between the SAMHD data and rectified CRC data (as shown in Figure 2) indicates the reliability of the proposed rectification method.

When using an unrectified CRC data set for the SEYAR model calibration, projections were significantly lower as compared to those obtained using the CRC data set, as shown in Figure 3 (a). On the other hand when using a rectified data set for calibration, projections were slightly higher than the CRC data, as shown in Figure 3(b). These findings suggest that forecasts obtained following model calibration with rectified data are less biased. Using the SAMHD data for calibration, the model’s average projected cases for July through mid-September 2020 were nearly in agreement with the CRC data (Figure 3 (c)).

It is worth noting that the distribution used in this study is measured from case data that were recorded during the early phase of the COVID-19 pandemic. Making the application of our proposed algorithm more suitable during the exponential growth phases of an outbreak.

Some possible limitations of the rectification algorithm deserve comment. First, the SAMHD data was available until July 7, 2020. After that date, changes in the reporting system made case tracing adjustments unavailable. Thereafter, the only baseline for comparison became CRC. Second, the measure of the delay distribution was based on data for Bexar County, Texas, (COVID-19 epidemiological event dates of confirmed cases) obtained from the San Antonio Metro Health District; based on our data, the optimal time that minimizes the reporting delay in CRC data was computationally estimated at 8 days. If there is evidence of a significantly higher delay in reporting in a different geographical region, the optimal data time should be measured for that location to reduce bias in the rectification.

The effect of reporting delays in data used for forecasting is often overlooked by many predictive models. Those models which utilize daily case counts often assume that the public case data are a faithful account of reality, which is hardly the case in practice as reporting delays in epidemic data remain inevitable. It is expected that the use of our methodological approach for rectifying and minimizing reporting delay when implementing prediction models could significantly improve the predictability of an epidemic.

## 5 Conclusion

To analyze the delays in case reporting to public health databases, we utilized both surveillance data and data obtained from public databases. We developed a method that rectifies such delays in public epidemiological data. Unlike the approaches reported in the literature, the rectification algorithm proposed here is premised on an exponential distribution that is measured from case tracing data with minimal assumptions. The method was shown to reliably account for the delays in reporting. The rectification of reporting delays in conjunction with our SEYAR prediction model seems promising for forecasting the actual number of daily cases. We stress the importance of capturing and publishing daily case counts as the data changes over time. This is particularly important as public databases such as the CRC do not currently update their daily case count records. This study has potential importance in terms of assessing the severity of a pandemic, particularly during its early stages. The results presented here also have the potential to aid in assessing the impact of possible control measures.

## Data Availability

All data produced in the present study are available upon reasonable request to the authors

